# Machine learning to classify left ventricular hypertrophy using ECG feature extraction by variational autoencoder

**DOI:** 10.1101/2024.10.14.24315460

**Authors:** Amulya Gupta, Christopher J. Harvey, Ashley DeBauge, Sumaiya Shomaji, Zijun Yao, Yongkuk Lee, Amit Noheria

**Affiliations:** Department of Cardiovascular Medicine, The University of Kansas Medical Center, Kansas City, Kansas; Department of Internal Medicine, Washington University School of Medicine, St. Louis, Missouri, USA; Department of Electrical Engineering and Computer Science, The University of Kansas, Lawrence, Kansas; Department of Biomedical Engineering, Wichita State University, Wichita, Kansas

**Keywords:** Left ventricular hypertrophy, LVH, machine learning, deep learning, artificial intelligence, electrocardiogram, ECG, variational autoencoder

## Abstract

**Background:** Traditional ECG criteria for left ventricular hypertrophy (LVH) have modest diagnostic yield.

**Objective:** Develop and validate machine learning models for LVH diagnosis from ECG.

**Methods:** ECG summary features (rate, intervals, axis), R-wave, S-wave and overall-QRS amplitudes, and QRS voltage-time integrals (VTI_QRS_) were extracted from 12-lead, vectorcardiographic X-Y-Z-lead, and 3D (L2 norm) representative-beat ECGs. Latent features (30 per ECG) were extracted using a variational autoencoder (trained on unselected >1 million ECGs) from X-Y-Z-lead representative-beat ECG signals. Logistic regression, random forest, light gradient boosted machine (LGBM), residual network (ResNet) and multilayer perceptron network (MLP) models using ECG features and sex, and a convolutional neural network (CNN) using ECG signals alone, were trained to predict LVH (left ventricular mass indexed in women >95 g/m^2^, men >115 g/m^2^) on 482,734 adult ECG-echocardiogram (within 45 days) pairs. ROC-AUCs for LVH classification are reported from a separate hold-out test set.

**Results:** In the test set (n=54,984), AUC for LVH classification was higher for ML models using ECG features (LGBM 0.794, MLP 0.793, ResNet 0.795) compared with the best individual ECG variable (VTI_QRS-Z_ 0.707), the best traditional criterion (Cornell voltage-duration product 0.716), and the CNN using ECG signals (0.788). Among patients without LVH who had a follow-up echocardiogram >1 (closest to 5) year later, LGBM false positives, compared to true negatives, had a 3.07 (95% CI 2.44, 3.86)-fold higher odds of developing future LVH (p<0.0001).

**Conclusions:** ML models are superior to traditional ECG criteria to classify LVH. Models trained on extracted ECG features, including latent variational autoencoder representations, can outperform CNN models directly trained on ECG signals.

## INTRODUCTION

<u>Left ventricular hypertrophy (LVH)</u> refers to increased left ventricular mass, characterized by an increase in left ventricular wall thickness and/or enlargement of the left ventricular cavity. This is often secondary to pathological or physiological stressors such as chronic hypertension, valvular heart disease, athletic training, or genetic conditions. LVH is associated with over a two-fold increase in cardiovascular morbidity and all-cause mortality (1). Early detection and initiation of pharmacological treatment and lifestyle modifications can improve outcomes (2).

<u>Transthoracic echocardiography</u> is the standard-of-care for the diagnosis of LVH. While non-invasive and widely available, echocardiography-based universal screening—even among high-risk populations, such as those with hypertension—is not cost-effective (3,4).

<u>Electrocardiography (ECG)</u> is an affordable, widely accessible, and frequently used diagnostic tool for cardiovascular screening. Often considered an extension of the cardiovascular physical examination, it is estimated that over 100-300 million ECGs are performed annually in the United States (5). Traditionally, LVH diagnosis via 12-lead ECG relies on voltage-based criteria, yet these show poor sensitivity, limiting their utility as standalone screening tools (6–8).

<u>Machine learning (ML)</u> can reduce reliance on human interpretation and yet increase the diagnostic accuracy of ECG (9,10). Several ECG-based ML models have been developed for detecting LVH, with varying sensitivities and specificities (11). Many of these studies use convolutional neural network (CNN) deep learning architectures to train models using ECG signals often with fewer than 10,000 training ECGs. Given that each 12-lead 10-second ECG signal at 500 Hz consists of 60,000 data points, using such a high-dimensionality input for ML training with a limited number of samples can result in overfitting and reduced generalizability (12–14). On the other hand, non-neural network ML architectures, such as logistic regression, random forest, gradient boosted machine, are not suited to use high-dimensional ECG signal data as input and are usually limited to using extracted ECG features, which may result in loss of diagnostic information (14).

To mitigate these limitations, while preserving the advantages of deep learning, we developed a <u>variational autoencoder (VAE)</u> that can encode 0.75-sec-representative-beat from either X-Y-Z-lead or root-mean-squared ECG into 30 variables (14–16). These VAE latent encodings retain the ECG morphological information and can reconstruct back the ECG signal with high fidelity. In this study, we aimed to train and test different ML models using extracted ECG features including the latent encodings or the ECG signal to classify LVH from the representative-beat ECG.

## METHODS

### Patient selection and data retrieval

An automated retrospective retrieval of records was performed from our clinical database at the University of Kansas Medical Center between May 2010 and January 2022 to search for ECG and echocardiogram performed on the same patient within 45 days of each other. Echocardiograms-ECG pairs with echocardiographic left ventricular mass index (LVMi) >95 g/m^2^ for females and >115 g/m^2^ for males were labelled as ‘LVH’ while rest of the pairs were assigned to the ‘no LVH’ group (14). The study was conducted with approval from the Institutional Review Board.

### Data extraction

ECGs were acquired from the Philips 12-lead ECG system. The 12-lead 1200-ms representative-beat signals along with standard features like heart rate, PR interval, etc. were extracted. Echocardiograms were standard clinical studies performed for clinical indications both as outpatient and inpatient evaluations. Individual echocardiogram numeric variables including diastolic measurements of left ventricular internal diameter (LVIDd), interventricular septum (IVSd) and posterior wall (PWd) from 2D parasternal long-axis view were extracted using a backend query in HERON (Healthcare Enterprise Repository for Ontological Narration), a search discovery tool that facilitates searches on various hospital electronic data sources (17,18). Left ventricular mass was calculated using the American Society of Echocardiography recommended formula: 0.8 × 1.04[(LVIDd + IVSd + PWd)^3^ and indexed to body surface area (19).

### ECG processing

The details of ECG processing performed using Python are provided in prior publications (14,20,21). In summary, vectorcardiographic X-Y-Z-lead ECGs were constructed from 12-lead ECGs using Kors’ matrix (22). L2 norm of these orthogonal leads signal was used to derive the 3D-ECG lead. Voltage-time integrals of QRS (VTI_QRS_) were obtained by the integration of the instantaneous voltage over the duration of QRS. R and S amplitudes were calculated as the maximum positive and negative deflections respectively of the QRS complex from the isoelectric line. Overall QRS amplitudes were calculated as the maximum absolute difference between the R and S deflections.

### Traditional Criteria and Univariable Models

Based on review of literature, we selected 5 widely used ECG-based LVH diagnostic criteria for comparison, i.e. Peguero-Lo Presti criteria (max S + S_v4_), Cornell voltage (R_avL_ + S_v3_), Cornell voltage-duration product (VDP), Sokolow-Lyon criteria (S_V1_ + max R _(V5 or V6)_), and Gubner-Ungerleider critera (R_I_ + S_III_) (23). We also selected ECG features for comparison namely QRS duration, Amplitude_QRS-3D_, and VTI_QRS-3D_ (20,21,23). The latter 2 were calculated off the QRS from the L2 norm/3D ECG. In addition, Amplitude_QRS_ and VTI_QRS_ in the X,Y, and Z projections were included.

### Variational Autoencoder

We trained a variational autoencoder (VAE) on 1.18 million unlabeled ECG signals to encode a 0.75-sec segment centered on the 1.2-sec representative beat ECG signal into 30 variables. The VAE has a dual neural network architecture with the encoder taking the ECG input and outputting 30 latent variables, and the decoder inputting the 30 latent variables and outputting the ECG signal. The network was rewarded in training to encode the signal such as to learn accurate reconstruction of the original signal from the latent variables alone. Our VAEs were able to reconstruct the original signal back from the latent variables with high fidelity (15,16,24). The X-Y-Z-lead representative-beat ECGs included in this study were processed using this VAE to generate latent encodings or variables.

### Model Input

For LVH diagnosis, these features were available for model training:

- Summary features like heart rate, PR interval, QRS duration, corrected QT interval (25), frontal plane QRS axis, etc.
- From 16 leads, each of 12-leads, 3 X-Y-Z-leads and 1 L2 norm/3D ECG, we obtained QRS amplitudes, VTI_QRS_, VTI_QRST_, R-wave amplitudes, S-wave amplitudes.
- 30 latent variables each from VAEs trained to reconstruct the X-Y-Z-lead and L2 norm/3D representative-lead ECGs.
- Sex

### Model Training and Testing

Approximately 10% of the medical record numbers in the dataset were withheld as the testing set, and the remainder were used for model training. We trained the following ML architectures on the training set: logistic regression, random forests, light gradient boosted machine (LGBM), residual neural network (ResNet), multilayered perceptron (MLP) and CNN. All the models were trained on the above mentioned features except for CNN, which was trained directly on the representative-beat X-Y-Z-lead ECG signal.

Sex was provided to the models as the definition of LVH is sex specific. The results are reported from the performance of the trained models in the holdout test set. We also report the models’ performance in 4 subgroups based on intraventricular conduction, QRS duration ≤120 ms, typical right bundle branch block (RBBB, QRS duration >120), typical left bundle branch block (LBBB, QRS duration >120 ms), and interventricular conduction delay (IVCD, QRS duration >120 ms but not meeting either RBBB or LBBB criteria). The American Heart Association-American College of Cardiology Foundation-Heart Rhythm Society criteria were used for classifying bundle branch blocks (26).

### Statistical analysis

Continuous variables are reported as mean ± standard deviation, and categorical variables as percentages. Comparisons were made using Student’s t-test for continuous variables and χ^2^-test for categorical variables. ECG processing and model training was performed in Python (version 3.12.7). Statistical analysis was conducted in R (version 4.4.1), and a two-tailed p-value of less than 0.05 was considered statistically significant.

## RESULTS

### Patient characteristics

A total of 537,718 ECG-echocardiogram pairs from 89,145 patients were included, with 237,592 (44.2%) pairs belonging to females. The mean age of the overall population of ECG-echocardiogram samples was 65.0 ± 15.4 years. This dataset was split into 90% training (*n =* 482,734) and 10% (*n =* 54,984) testing sets. In the training set, 70,659 (33.1%) of the female samples and 77,777 (28.9%) male samples had echocardiographic LVH. In the testing set, 8,051 (33.6%) female samples and 8,793 (28.4%) male samples had LVH. The detailed distributions of the ECG and echocardiographic variables in the testing set are shown in **Table 1** and for the training set in **Supplementary Table 1**. As shown in **Figure 1**, the testing samples were divided into 4 subgroups i.e. narrow QRS <120 ms (*n* = 39,936; 72.6%), typical RBBB (*n* = 6,731; 12.2%), typical LBBB (*n* = 6,372; 11.6%), and IVCD (*n* = 1,945; 3.5%).

**Figure 1.**
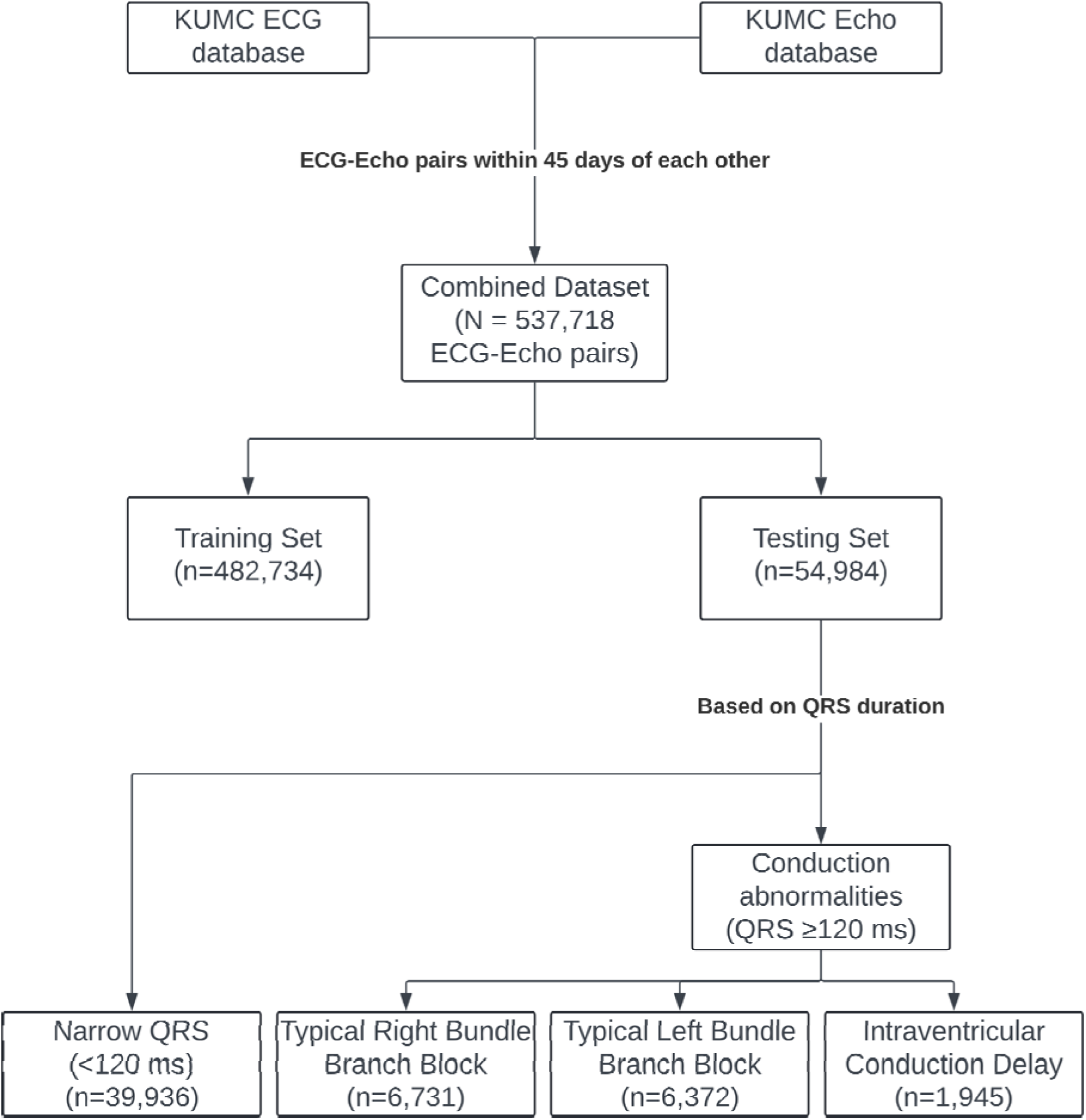
Data pipeline for model development and testing.

**Table 1.**
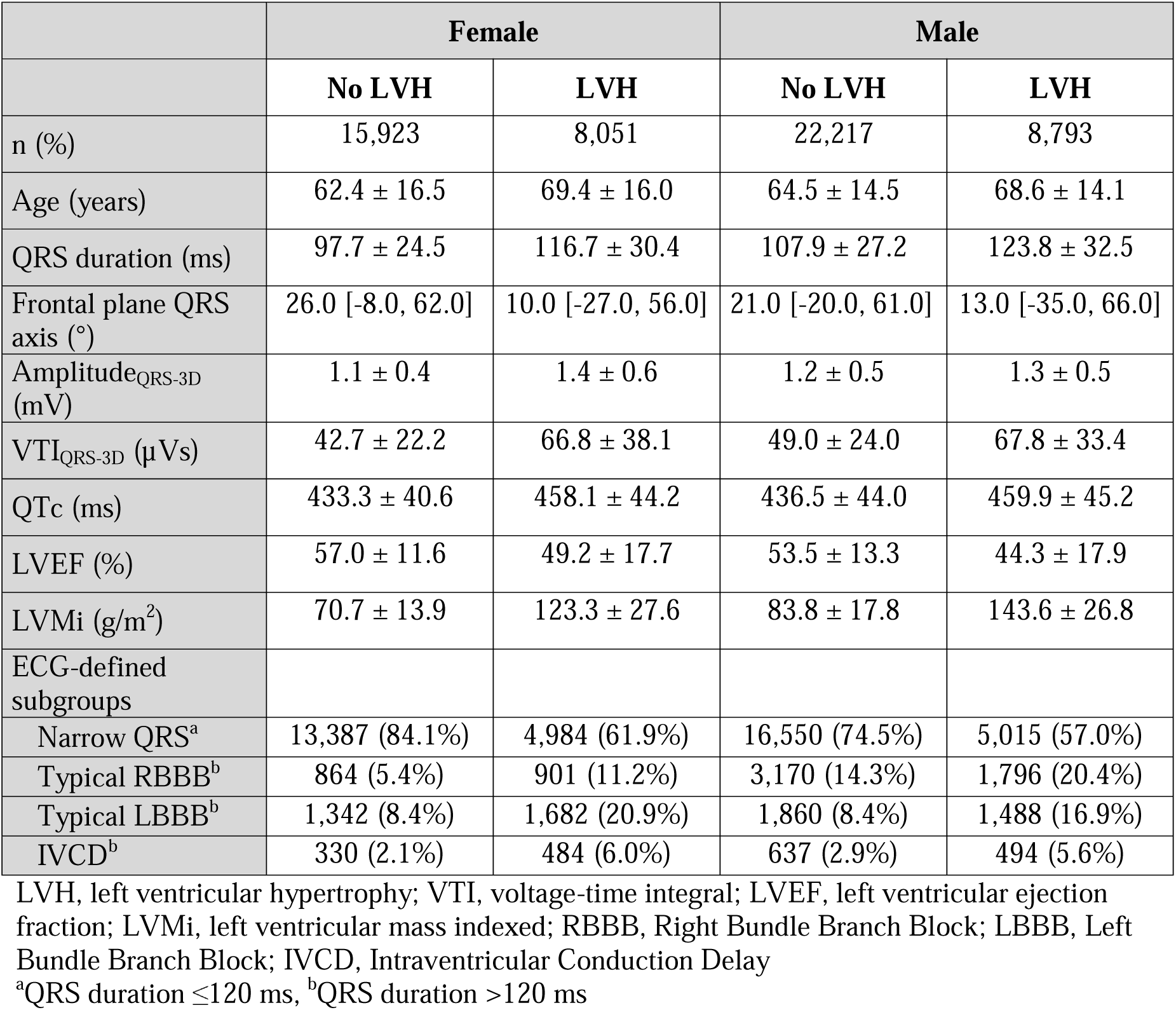
Patient characteristics of the validation set.

|  | Female |  | Male |  |
| --- | --- | --- | --- | --- |
|  | No LVH | LVH | No LVH | LVH |
| n (%) | 15,923 | 8,051 | 22,217 | 8,793 |
| Age (years) | 62.4 ± 16.5 | 69.4 ± 16.0 | 64.5 ± 14.5 | 68.6 ± 14.1 |
| QRS duration (ms) | 97.7 ± 24.5 | 116.7 ± 30.4 | 107.9 ± 27.2 | 123.8 ± 32.5 |
| Frontal plane QRS axis (°) | 26.0 [-8.0, 62.0] | 10.0 [-27.0, 56.0] | 21.0 [-20.0, 61.0] | 13.0 [-35.0, 66.0] |
| Amplitude<sub>QRS-3D</sub> (mV) | 1.1 ± 0.4 | 1.4 ± 0.6 | 1.2 ± 0.5 | 1.3 ± 0.5 |
| VTI<sub>QRS-3D</sub> (μVs) | 42.7 ± 22.2 | 66.8 ± 38.1 | 49.0 ± 24.0 | 67.8 ± 33.4 |
| QTc (ms) | 433.3 ± 40.6 | 458.1 ± 44.2 | 436.5 ± 44.0 | 459.9 ± 45.2 |
| LVEF (%) | 57.0 ± 11.6 | 49.2 ± 17.7 | 53.5 ± 13.3 | 44.3 ± 17.9 |
| LVMi (g/m<sup>2</sup>) | 70.7 ± 13.9 | 123.3 ± 27.6 | 83.8 ± 17.8 | 143.6 ± 26.8 |
| ECG-defined subgroups |  |  |  |  |
| Narrow QRS<sup>a</sup> | 13,387 (84.1%) | 4,984 (61.9%) | 16,550 (74.5%) | 5,015 (57.0%) |
| Typical RBBB<sup>b</sup> | 864 (5.4%) | 901 (11.2%) | 3,170 (14.3%) | 1,796 (20.4%) |
| Typical LBBB<sup>b</sup> | 1,342 (8.4%) | 1,682 (20.9%) | 1,860 (8.4%) | 1,488 (16.9%) |
| IVCD<sup>b</sup> | 330 (2.1%) | 484 (6.0%) | 637 (2.9%) | 494 (5.6%) |

LVH, left ventricular hypertrophy; VTI, voltage-time integral; LVEF, left ventricular ejection fraction; LVMi, left ventricular mass indexed; RBBB, Right Bundle Branch Block; LBBB, Left Bundle Branch Block; IVCD, Intraventricular Conduction Delay
<sup>a</sup>QRS duration ≤120 ms, <sup>b</sup>QRS duration >120 ms

### LVH classification models

The testing set performance of the univariable models, traditional criteria and ML models is summarized in **Table 2**. Model performance statistics in QRS morphological subgroups are available **Supplementary Table 2A-D**.

**Table 2.** Model performance for LVH prediction in the entire validation set. Area under receiver-operating characteristic curve (AUC) and sensitivity at specificity fixed at 0.75 are provided.

| Validation set | Combined<br>(n=54,984) |  | Females<br>(n=23,974) |  | Males<br>(n=31,010) |  |
| --- | --- | --- | --- | --- | --- | --- |
|  | AUC | Sensitivity* | AUC | Sensitivity* | AUC | Sensitivity* |
| **<u>Univariable models<sup>a</sup></u>** |  |  |  |  |  |  |
| ▪ QRS duration | 0.672 | 0.454 | 0.705 | 0.469 | 0.659 | 0.46 |
| ▪ Amplitude<sub>QRS-3D</sub> | 0.594 | 0.388 | 0.610 | 0.416 | 0.580 | 0.362 |
| ▪ Amplitude<sub>QRS-X</sub> | 0.514 | 0.308 | 0.518 | 0.324 | 0.517 | 0.304 |
| ▪ Amplitude<sub>QRS-Y</sub> | 0.523 | 0.281 | 0.523 | 0.275 | 0.514 | 0.276 |
| ▪ Amplitude<sub>QRS-Z</sub> | 0.661 | 0.478 | 0.686 | 0.526 | 0.643 | 0.445 |
| ▪ VTI<sub>QRS-3D</sub> | 0.704 | 0.535 | 0.721 | 0.569 | 0.699 | 0.518 |
| ▪ VTI<sub>QRS-X</sub> | 0.622 | 0.442 | 0.638 | 0.468 | 0.621 | 0.439 |
| ▪ VTI<sub>QRS-Y</sub> | 0.615 | 0.408 | 0.628 | 0.422 | 0.603 | 0.392 |
| ▪ VTI<sub>QRS-Z</sub> | 0.707 | 0.533 | 0.733 | 0.581 | 0.692 | 0.500 |
| **<u>Traditional criteria</u>** |  |  |  |  |  |  |
| ▪ Peguero-Lo Presti voltage (max S + S<sub>V4</sub>) | 0.694 | 0.523 | 0.721 | 0.570 | 0.677 | 0.497 |
| ▪ Cornell voltage (R<sub>avL</sub> + S<sub>V3</sub>) | 0.670 | 0.493 | 0.704 | 0.545 | 0.646 | 0.459 |
| ▪ Cornell VDP | 0.716 | 0.555 | 0.747 | 0.607 | 0.689 | 0.507 |
| ▪ Sokolow-Lyon voltage (S<sub>V1</sub> + max R (V5 or V6)) | 0.545 | 0.345 | 0.563 | 0.375 | 0.529 | 0.322 |
| ▪ Gubner-Ungerleider voltage (R<sub>I</sub> + S<sub>III</sub>) | 0.566 | 0.358 | 0.610 | 0.407 | 0.522 | 0.310 |
| **<u>ML models</u>** |  |  |  |  |  |  |
| ▪ Logistic regression<sup>b</sup> | 0.783 | 0.662 | 0.792 | 0.684 | 0.773 | 0.639 |
| ▪ Random forest<sup>b</sup> | 0.779 | 0.656 | 0.794 | 0.687 | 0.769 | 0.634 |
| ▪ Light gradient boosted machine<sup>b</sup> | 0.794 | 0.684 | 0.812 | 0.719 | 0.786 | 0.666 |
| ▪ Residual network<sup>b</sup> | 0.795 | 0.686 | 0.804 | 0.709 | 0.785 | 0.663 |
| ▪ Multilayered perceptron network<sup>b</sup> | 0.793 | 0.681 | 0.802 | 0.700 | 0.784 | 0.661 |
| ▪ Convolutional neural network<sup>c</sup> | 0.788 | 0.673 | 0.803 | 0.702 | 0.782 | 0.661 |

VDP, voltage-duration product; ML, machine learning
\* At specificity 0.75
<sup>a</sup> Logistic regressions.
<sup>b</sup> Input of ECG statistics like QRS duration, heart rate etc. and 30 variational autoencoder latent variables from ECG representative beat and sex.
<sup>c</sup> Input of representative-beat ECG signal (X, Y, Z leads)

### Univariable models

Amongst the linear univariable models, VTI_QRS-Z_ was the best predictor of LVH in the overall population, with an AUC 0.707 which was closely followed by VTI_QRS-3D_ with AUC 0.704. Furthermore, VTI_QRS-Z_ demonstrated the highest performance in the narrow QRS subgroup (AUC 0.704), whereas VTI_QRS-3D_ performed best in the typical RBBB subgroup (AUC 0.683). The best performing criteria in typical LBBB was Amplitude_QRS-3D_ (0.655) and in IVCD was QRS duration (0.677).

### Traditional criteria

Overall, the performance of traditional ECG criteria for predicting LVH was fair, with AUCs ranging from 0.545 to 0.716. Cornell VDP was the best performing criteria in all groups except typical LBBB (AUC overall 0.716; narrow QRS 0.713, typical RBBB 0.678, IVCD 0.694). In the typical LBBB subgroup, Peguero-Lo Presti criteria (0.651) performed the best. In general, these criteria performed better in females as compared to males.

### ML Models

All ML models outperformed the traditional criteria and univariate models.

LGBM (AUC 0.794), MLP (0.793) and ResNet (0.795), which were trained on ECG features including VAE latent encodings and sex, were the best performing models in the overall population. The CNN model, which was trained on the raw ECG signal alone, demonstrated an AUC 0.788. The ROC curves, separately for females and males, for the top 4 ML models vis-à-vis the best univariable and best traditional criteria are plotted in **Figure 2**.

**Figure 2.**
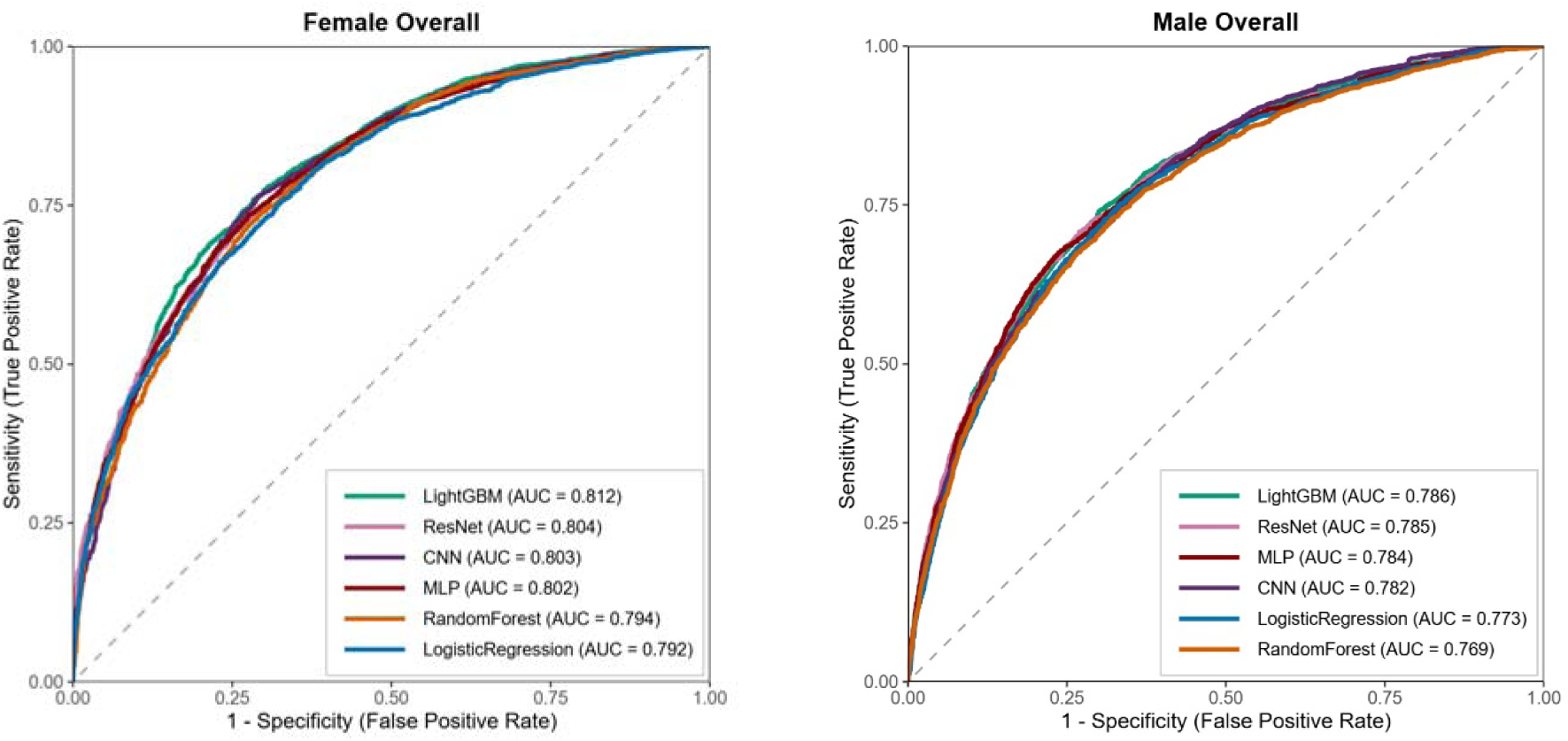
ROC curves for machine learning models from the entire testing set for males (left panel) and females (right panel).

ROC curves for VTI_QRS-3D_, Cornell VDP, and LGBM for overall test set and each of the 4 subgroups separately for females and males are shown in **Figure 3 and 4**.

**Figure 3.**
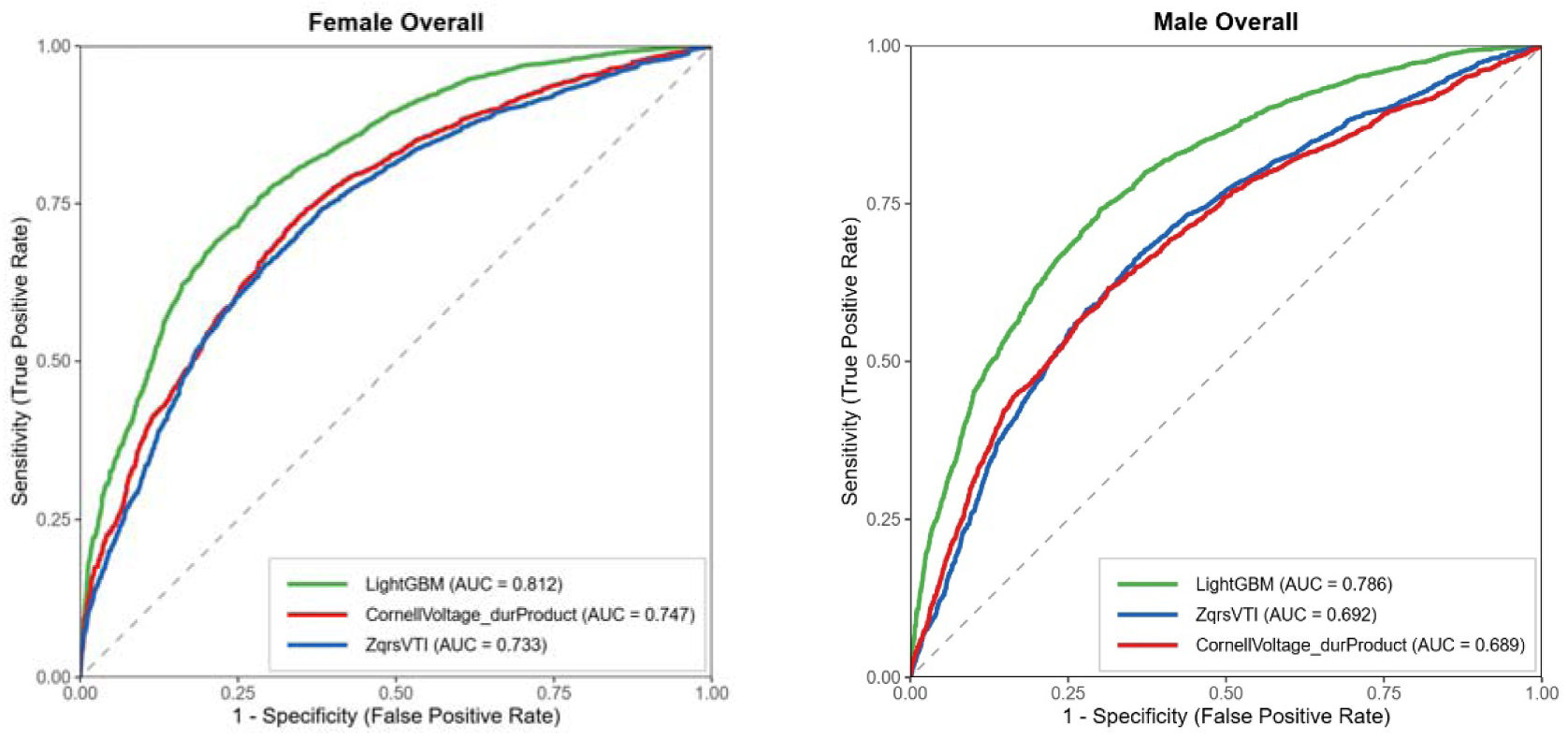
ROC curves for best peforming LVH models – univariate (VTI¬QRS-Z), traditional criteria (Cornell VDP), and machine learning (LGBM) from the entire testing set for females (left) and males (right).

**Figure 4.**
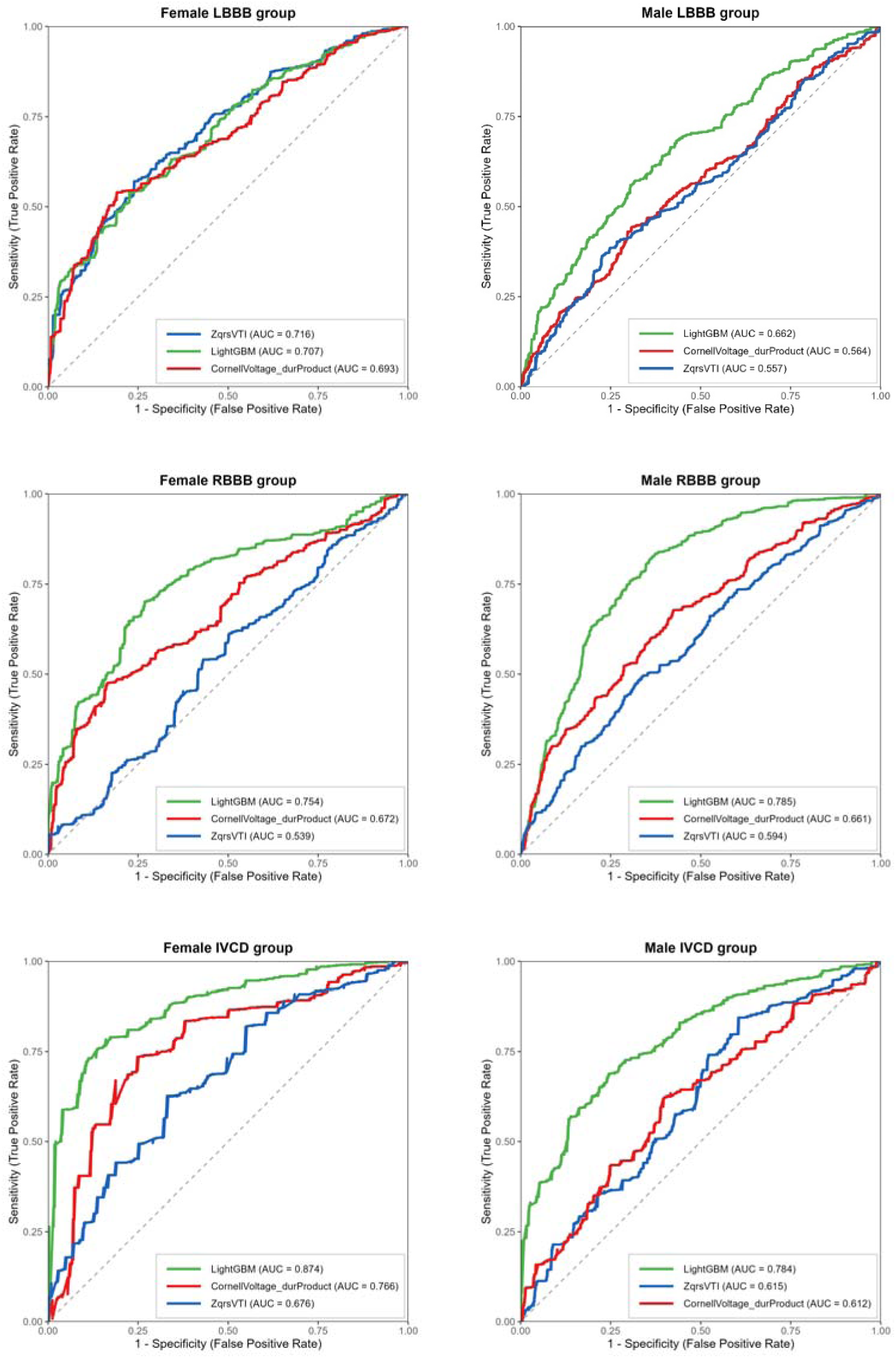
ROC curves for best peforming LVH models – univariate (VTI¬QRS-Z), traditional criteria (Cornell VDP), and machine learning (LGBM) from the wide QRS subgroups (> 120 ms) for females (left) and males (right).

### Linear analysis of LGBM prediction probabilities

LVMi was plotted against the prediction probabilities output generated by LGBM model for females and males as shown in **Figure 5**. A strong linear trend between prediction probabilities and LVMi can be noted for both females and males (respectively R^2^ 0.34 and 0.26; correlation coefficient 0.582 and 0.508).

**Figure 5.**
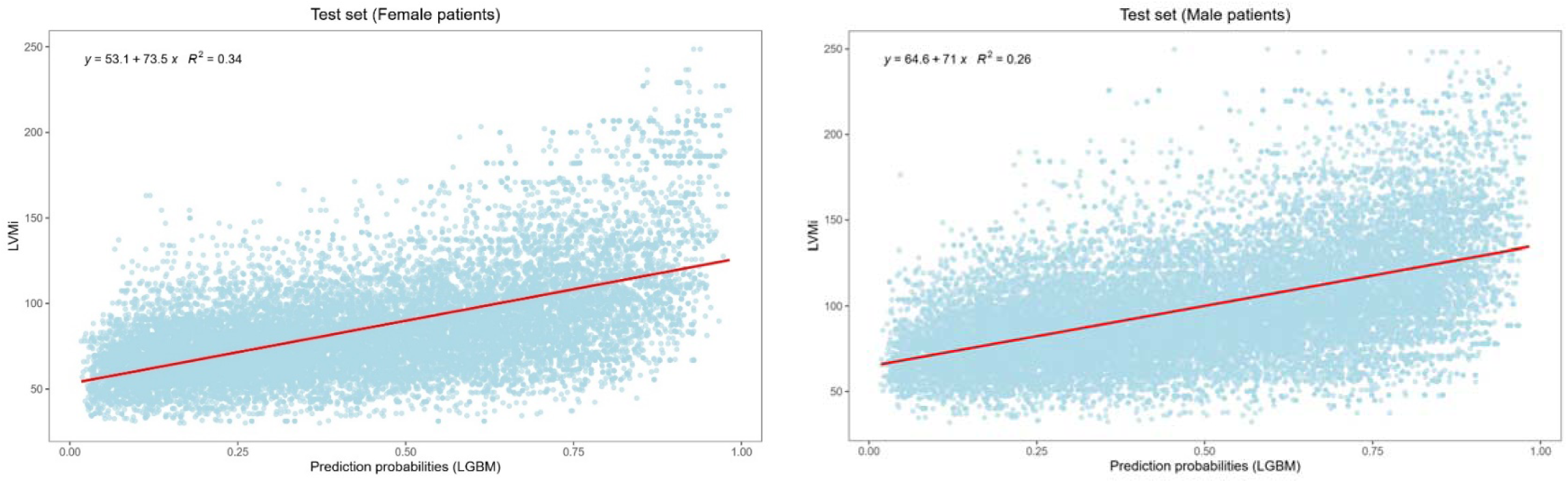
Scatterplots of echocardiographic left ventricular mass indexed (LVMi) plotted against prediction probabilities from the LGBM model for females (left panel) and males (right panel).

### Longitudinal analysis of LVH negatives

Among false positives and true negatives produced by the LGBM model in the validation set, we searched for the ECG-echocardiogram pairs where a follow-up echocardiogram >1 year and closest to 5 years later was available for further analysis. We used a 2×2 table to compare the development of LVH in 612 false-positive as compared to the 1,543 true-negative samples. On mean follow-up of 3.3 ± 1.7 years, 189/612 (30.9%) patients in false-positive group, and 196/1543 (12.7%) patients in true-negative group developed LVH. The odds ratio for development of LVH was 3.07 (95% CI 2.44, 3.86, p<0.0001) in false-positives compared to true-negatives from the LGBM model (**Table 3**).

**Table 3.**
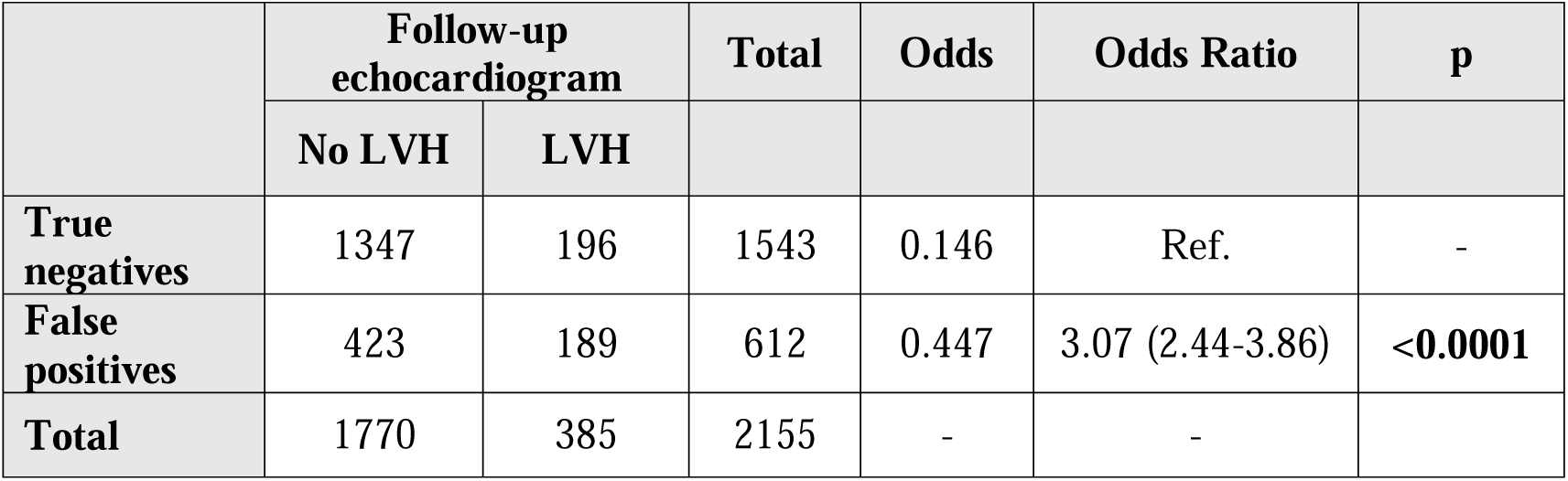
Comparison between presence of LVH on subsequent echocardiogram (>1 year and closest to 5 years after index echocardiogram) in false positives versus true negatives of LVH LGBM model in testing set.

## DISCUSSION

To the best of our knowledge, this study represents the largest evaluation of ECG criteria and ML models for LVH prediction to date. We have applied the innovative framework of using DL-based latent space ECG encodings for building ML models, which allows simpler models to make accurate predictions without overfitting.

### Salient findings

First, traditional ECG-based criteria demonstrated moderate utility in diagnosing LVH, with the Cornell VDP showing the highest discrimination among them (AUC 0.716). Second, univariable criteria including VTI_QRS-Z_ and VTI_QRS-3D_ were at par with traditional criteria for the diagnosis of LVH, with VTI_QRS-Z_ achieving the best overall results (AUC 0.707). Third, our ML models outperform both traditional and univariable models, with LGBM, ResNet, and MLP demonstrating the highest performance (AUC 0.793-0.795). Last, the performance of traditional, univariable, and ML models vary across sex and QRS morphologies. Furthermore, the LGBM model trained on ECG latent encodings and features captured the underlying trend of cardiac mass, not just the LVH labels, showing strong correlation with LVMi and predicting future development of LVH.

### Univariable models

Previous studies have demonstrated the utility of linear univariable predictors of LVH, such as QRS duration and area (21,27). In our analysis, we evaluated QRS duration and vectorcardiographic amplitudes/voltage-time integrals for predicting LVH across various subgroups. Among them, VTI_QRS-Z_ and VTI_QRS-3D_ emerged as the best overall criteria. Similar to Cornell VDP, both voltage and duration components of the QRS complex are incorporated in these criteria. Prolongation of the QRS duration in LVH is likely attributable to delays in ventricular activation associated with increased wall thickness and frequent conduction abnormalities. Furthermore, since VTI_QRS-3D_ is calculated from the reconstructed 3D-orthogonal leads, it ostensibly captures the sum of net surface potentials produced by the ventricular depolarization wavefront in a cardiac cycle. The absolute value of surface potentials, especially in Z-axis (sagittal axis) are likely to be increased due to LVH as the mean depolarization vector shifts leftwards and posteriorly (28). Consequently, VTI_QRS-Z_ and VTI_QRS-3D_ are likely to be increased in LVH and could be explored as a simple univariable predictor of LVH.

### Traditional ECG criteria

As demonstrated in previous studies, our analysis reaffirmed the modest-to-fair discrimination of LVH offered by standard electrocardiographic criteria using a large dataset (29,30). Unlike other voltage-based rules, Cornell VDP, which emerged as the best overall criterion, accounts for both QRS voltage and duration in its calculation, both of which are affected in LVH (31). In the subset of ECGs with LBBB, Peguero-Lo Presti criteria performed better than Cornell VDP. Although the difference in performance was marginal, if this trend is physiological, it could be explained by obfuscation of LVH-related changes in QRS duration due to QRS prolongation inherent to conduction delays in LBBB. However, this cannot be verified in our study. Additionally, compared to the combined population, individual criteria generally performed better in females and males separately. This underscores the importance of using different cut-off values for females and males, recognizing the sex-based differences in ECGs and definition of LVH (29,30).

### ML models

We tested several ML architectures for LVH prediction, including simple models (LR), tree-based models (RF, LGBM), and neural networks (ResNet, MLP, and CNN). The best overall performance was shown by LGBM, ResNet, and MLP with AUCs ∼0.79.

Performance declined across all models in the subgroups with conduction abnormalities. Interestingly, these models, despite only using the summary ECG features and VAE encodings, performed slightly better than the CNN model, which had access to the raw signal data. The use of the LGBM architecture for such classification tasks is pragmatic for two main reasons. First, as an implementation of gradient boosting decision trees, it enables efficient model training with relatively low computational resources. Second, its decision-making process is inherently interpretable, as feature importance can be evaluated using metrics like split count and gain.

We further evaluated the interpretability and physiological relevance of the LGBM model.

First, we plotted the prediction probabilities from this model against LVMi, which showed a strong linear positive correlation, suggesting that the model captures meaningful physiological patterns rather than artificial class boundaries. Second, we analyzed the false positives produced by this model for future development of LVH, finding that the false positives had 3 times the odds of developing LVH in the future compared to true negatives. This indicates that the model may capture underlying ECG abnormalities even before patients meet the criteria for overt LVH diagnosis.

### Previous literature

In a recently published study from China, Zhu et al. used a large dataset comprising of over 90,000 ECGs to create deep learning multilabel classifier algorithms. They achieved AUCs ranging from 0.78-0.92 using their 12-lead model and showed that a reduced 4-lead model using lead I, aVR, V1 and V5 had equivalent performance (32). In a Taiwanese study, Liu et al. developed a deep learning model for predicting LVH using approximately 23,000 training samples (33). They achieved high AUCs ranging from 0.83-0.89 across different validation sets. However, the definition of LVH used in this study was different, using LV mass >186 g for females and >258 g for males. In a South Korean study, Kwon et al. developed an ensemble deep neural network + CNN model using approximately 36,000 training samples, combining information from ECG signal, ECG features, and patient demographics (34). While using higher cut-off values for LVMi (109 g/m^2^ females and 132 g/m^2^ males), their model achieved AUCs ranging from 0.87-0.88 in validation sets.

In a study from Massachusetts General Hospital, Haimovich et al. create ML models for predicting LVH in specific disease populations like cardiac amyloidosis, hypertrophic cardiomyopathy, aortic stenosis, and others using a total of 34,258 training samples (35). Similar to our approach, they used a pretrained deep learning model to produce latent encodings and trained a simpler classifier for LVH classification although they used full 10-second ECG signal instead of representative beat ECG. Their model achieved AUCs ranging from 0.69 to 0.96 in various subgroups. Khurshid et al. used data from the UK Biobank to create a CNN model trained on 32,000 samples and achieved AUCs ranging from 0.62 to 0.65 in predicting LVH. Owing to heterogeneity in study populations, data structures, and labels for LVH, it is difficult to evaluate the performance of models across studies. Nonetheless, the AUCs attained by ML models in our study are comparable to previous work.

### Limitations

Our work is best understood in the context of its limitations. Both training and testing sets for the models were from a single center, and these models might have sub-optimal performance when generalized to other datasets. The inclusion of in-hospital ECGs may have biased our dataset towards sicker patients, who often get multiple ECGs in each hospital stay. Further, since the median beat ECGs were derived from a proprietary system, additional steps may be required in processing ECGs from other systems. Additionally, to calculate ECG parameters for traditional criteria and univariate models, automated feature extraction was done, which might not be as accurate as expert-created labels.

## CONCLUSIONS

Traditional voltage-based ECG criteria demonstrate only modest performance in detecting LVH. Simple univariable models, especially VTI_QRS-Z_, may perform at par with traditional criteria.

Regardless, ML techniques can significantly enhance the accuracy of ECG-based diagnosis of LVH over both traditional voltage-based criteria and univariable models. Our findings highlight the utility of dimensionality reduction via variational autoencoders, which enables the application of non-deep learning ML models to high-dimensional ECG data without compromising performance. These approaches offer both interpretability and scalability, suggesting potential for future clinical integration. Further external validation and testing is needed for clinical utilization of these ML models.

## Data Availability

The data supporting findings of this study were obtained from our institutional database that contains identifiable patient information. Access to the data is restricted and subject to approval by the institutional review board. Researchers interested in accessing the data may contact the corresponding author for information about the necessary procedures and approvals required.

## Abbreviations

ECG: electrocardiogram
LVH: left ventricular hypertrophy
ML: machine learning
AI: artificial intelligence
MLP: multilayered perceptron
LGBM: light gradient-boosting machine
AUC: area under the receiver operator characteristic curve
VAE: variational Autoencoder
LVMi: left ventricular mass indexed

