## Supplementary File for "Machine learning to classify left ventricular hypertrophy using ECG feature extraction by variational autoencoder"

**Supplementary table 1.** Patient characteristics of the training set.

|  | **Female** | | **Male** | |
| --- | --- | --- | --- | --- |
|  | **No LVH** | **LVH** | **No LVH** | **LVH** |
| n (%) | 142,959 | 70,659 | 191,339 | 77,777 |
| Age (years) | 62.6 ± 16.5 | 68.7 ± 14.7 | 64.1 ± 15.1 | 68.2 ± 13.4 |
| QRS duration (ms) | 96.8 ± 22.8 | 110.9 ± 28.8 | 106.3 ± 26.2 | 122.9 ± 31.0 |
| Frontal plane QRS axis (°) | 26.0 [-8.0, 59.0] | 8.0 [-27.0, 49.0] | 21.0 [-17.0, 59.0] | 5.0 [-35.0, 54.0] |
| Amplitude_QRS-3D_ (mV) | 1.1 ± 0.4 | 1.3 ± 0.5 | 1.2 ± 0.4 | 1.3 ± 0.5 |
| VTI_QRS-3D_ (µVs) | 42.1 ± 21.7 | 58.7 ± 32.0 | 48.7 ± 24.6 | 66.9 ± 33.4 |
| QTc (ms) | 434.2 ± 40.2 | 453.3 ± 46.2 | 436.1 ± 42.7 | 458.7 ± 45.8 |
| LVEF (%) | 57.3 ± 11.4 | 50.3 ± 16.3 | 53.0 ± 13.5 | 43.6 ± 21.0 |
| LVMi (g/m^2^) | 71.0 ± 14.0 | 120.2 ± 22.6 | 84.2 ± 17.6 | 141.1 ± 23.8 |
| ECG-defined subgroups |  |  |  |  |
| Narrow QRS^a^ | 122,078 (85.4%) | 48,433 (68.5%) | 147,528 (77.1%) | 43,614 (56.1%) |
| Typical RBBB^b^ | 7,668 (5.4%) | 7,329 (10.4%) | 21,847 (11.4%) | 14,673 (18.9%) |
| Typical LBBB^b^ | 10,483 (7.3%) | 12,090 (17.1%) | 16,663 (8.7%) | 14,777 (19.0%) |
| IVCD^b^ | 2,730 (1.9%) | 2,807 (4.0%) | 5,301 (2.8%) | 4,713 (6.1%) |

LVH, left ventricular hypertrophy; VTI, voltage-time integral; LVEF, left ventricular ejection fraction; LVMi, left ventricular mass indexed; RBBB, Right Bundle Branch Block; LBBB, Left Bundle Branch Block; IVCD, Intraventricular Conduction Delay.

^a^QRS duration ≤120 ms, ^b^QRS duration >120 ms

**Supplementary table 2A.** Model performance for LVH prediction in population **with narrow QRS duration** (≤120 ms) in the test set.

| **Validation set** | **Combined**  **(n=39,936)** | | **Females**  **(n=18,371)** | | **Males**  **(n=21,565)** | |
| --- | --- | --- | --- | --- | --- | --- |
|  | **AUROC** | **Sensitivity*** | **AUROC** | **Sensitivity*** | **AUROC** | **Sensitivity*** |
| ***Univariable models^a^*** |  |  |  |  |  |  |
| - QRS duration | 0.647 | 0.414 | 0.668 | 0.437 | 0.645 | 0.412 |
| - Amplitude_QRS-3D_ | 0.592 | 0.385 | 0.587 | 0.386 | 0.597 | 0.387 |
| - Amplitude_QRS-X_ | 0.548 | 0.347 | 0.551 | 0.363 | 0.552 | 0.345 |
| - Amplitude_QRS-Y_ | 0.522 | 0.284 | 0.490 | 0.232 | 0.528 | 0.292 |
| - Amplitude_QRS-Z_ | 0.668 | 0.484 | 0.673 | 0.496 | 0.672 | 0.488 |
| - VTI_QRS-3D_ | 0.672 | 0.500 | 0.677 | 0.504 | 0.683 | 0.518 |
| - VTI_QRS-X_ | 0.594 | 0.405 | 0.602 | 0.416 | 0.598 | 0.413 |
| - VTI_QRS-Y_ | 0.569 | 0.342 | 0.557 | 0.320 | 0.577 | 0.359 |
| - VTI_QRS-Z_ | 0.704 | 0.537 | 0.717 | 0.554 | 0.702 | 0.537 |
| ***Traditional criteria*** |  |  |  |  |  |  |
| - Peguero-Lo Presti voltage (max S + S_V4_) | 0.693 | 0.523 | 0.699 | 0.53 | 0.704 | 0.542 |
| - Cornell voltage (R_avL_ + S_V3_) | 0.679 | 0.505 | 0.694 | 0.523 | 0.680 | 0.508 |
| - Cornell VDP | 0.713 | 0.553 | 0.727 | 0.568 | 0.698 | 0.536 |
| - Sokolow-Lyon voltage (S_V1_ + max R (V5 or V6)) | 0.587 | 0.391 | 0.583 | 0.396 | 0.594 | 0.392 |
| - Gubner-Ungerleider voltage (R_I_ + S_III_) | 0.566 | 0.363 | 0.591 | 0.388 | 0.540 | 0.335 |
| ***ML models*** |  |  |  |  |  |  |
| - Logistic regression^b^ | 0.776 | 0.654 | 0.778 | 0.662 | 0.772 | 0.645 |
| - Random forest^b^ | 0.771 | 0.643 | 0.785 | 0.663 | 0.759 | 0.628 |
| - Light gradient boosted machine^b^ | 0.790 | 0.677 | 0.805 | 0.7 | 0.78 | 0.664 |
| - Residual network^b^ | 0.788 | 0.675 | 0.792 | 0.685 | 0.782 | 0.665 |
| - Multilayered perceptron network^b^ | 0.785 | 0.668 | 0.79 | 0.677 | 0.777 | 0.659 |
| - Convolutional neural network^c^ | 0.782 | 0.663 | 0.792 | 0.68 | 0.778 | 0.658 |

VDP, voltage-duration product; ML, machine learning

* At specificity 0.75

^a^ Logistic regressions

^b^ Input of ECG statistics like QRS duration, heart rate etc. and 30 variational autoencoder latent variables from ECG representative beat and sex

^c^ Input of representative-beat ECG signal (X, Y, Z leads)

**Supplementary table 2B.** Model performance for LVH prediction in population with typical right bundle branch block (QRS duration >120 ms) validation set.

| **Validation set** | **Combined**  **(n=6,731)** | | **Females**  **(n=1,765)** | | **Males**  **(n=4,966)** | |
| --- | --- | --- | --- | --- | --- | --- |
|  | **AUROC** | **Sensitivity*** | **AUROC** | **Sensitivity*** | **AUROC** | **Sensitivity*** |
| ***Univariable models^a^*** |  |  |  |  |  |  |
| - QRS duration | 0.619 | 0.383 | 0.569 | 0.276 | 0.647 | 0.422 |
| - Amplitude_QRS-3D_ | 0.616 | 0.396 | 0.647 | 0.452 | 0.598 | 0.354 |
| - Amplitude_QRS-X_ | 0.571 | 0.361 | 0.640 | 0.444 | 0.548 | 0.335 |
| - Amplitude_QRS-Y_ | 0.541 | 0.316 | 0.589 | 0.382 | 0.502 | 0.260 |
| - Amplitude_QRS-Z_ | 0.586 | 0.373 | 0.572 | 0.369 | 0.595 | 0.372 |
| - VTI_QRS-3D_ | 0.683 | 0.504 | 0.684 | 0.509 | 0.679 | 0.494 |
| - VTI_QRS-X_ | 0.661 | 0.485 | 0.706 | 0.553 | 0.648 | 0.467 |
| - VTI_QRS-Y_ | 0.605 | 0.396 | 0.651 | 0.443 | 0.572 | 0.352 |
| - VTI_QRS-Z_ | 0.578 | 0.362 | 0.539 | 0.305 | 0.594 | 0.383 |
| ***Traditional criteria*** |  |  |  |  |  |  |
| - Peguero-Lo Presti voltage (max S + S_V4_) | 0.668 | 0.486 | 0.688 | 0.502 | 0.664 | 0.484 |
| - Cornell voltage (R_avL_ + S_V3_) | 0.650 | 0.456 | 0.683 | 0.509 | 0.642 | 0.444 |
| - Cornell VDP | 0.678 | 0.503 | 0.672 | 0.503 | 0.661 | 0.472 |
| - Sokolow-Lyon voltage (S_V1_ + max R (V5 or V6)) | 0.501 | 0.263 | 0.534 | 0.316 | 0.515 | 0.279 |
| - Gubner-Ungerleider voltage (R_I_ + S_III_) | 0.533 | 0.303 | 0.648 | 0.455 | 0.520 | 0.282 |
| ***ML models*** |  |  |  |  |  |  |
| - Logistic regression^b^ | 0.738 | 0.586 | 0.675 | 0.521 | 0.755 | 0.603 |
| - Random forest^b^ | 0.765 | 0.622 | 0.731 | 0.597 | 0.776 | 0.632 |
| - Light gradient boosted machine^b^ | 0.773 | 0.632 | 0.754 | 0.629 | 0.785 | 0.649 |
| - Residual network^b^ | 0.772 | 0.636 | 0.749 | 0.609 | 0.778 | 0.639 |
| - Multilayered perceptron network^b^ | 0.773 | 0.636 | 0.735 | 0.584 | 0.784 | 0.648 |
| - Convolutional neural network^c^ | 0.766 | 0.631 | 0.767 | 0.634 | 0.771 | 0.644 |

VDP, voltage-duration product; ML, machine learning

* At specificity 0.75

^a^ Logistic regressions

^b^ Input of ECG statistics like QRS duration, heart rate etc. and 30 variational autoencoder latent variables from ECG representative beat and sex

^c^ Input of representative-beat ECG signal (X, Y, Z leads)

**Supplementary table 2C.** Model performance for LVH prediction in population with typical left bundle branch block (QRS duration >120 ms) validation set.

| **Validation set** | **Combined**  **(n=6,372)** | | **Females**  **(n=3,024)** | | **Males**  **(n=3,348)** | |
| --- | --- | --- | --- | --- | --- | --- |
|  | **AUROC** | **Sensitivity*** | **AUROC** | **Sensitivity*** | **AUROC** | **Sensitivity*** |
| ***Univariable models^a^*** |  |  |  |  |  |  |
| - QRS duration | 0.550 | 0.284 | 0.601 | 0.293 | 0.530 | 0.289 |
| - Amplitude_QRS-3D_ | 0.655 | 0.468 | 0.732 | 0.592 | 0.579 | 0.344 |
| - Amplitude_QRS-X_ | 0.584 | 0.386 | 0.582 | 0.399 | 0.590 | 0.377 |
| - Amplitude_QRS-Y_ | 0.578 | 0.321 | 0.617 | 0.358 | 0.535 | 0.280 |
| - Amplitude_QRS-Z_ | 0.652 | 0.461 | 0.726 | 0.585 | 0.584 | 0.347 |
| - VTI_QRS-3D_ | 0.642 | 0.445 | 0.718 | 0.577 | 0.576 | 0.343 |
| - VTI_QRS-X_ | 0.593 | 0.386 | 0.591 | 0.396 | 0.594 | 0.373 |
| - VTI_QRS-Y_ | 0.587 | 0.328 | 0.624 | 0.361 | 0.545 | 0.293 |
| - VTI_QRS-Z_ | 0.630 | 0.424 | 0.716 | 0.566 | 0.557 | 0.321 |
| ***Traditional criteria*** |  |  |  |  |  |  |
| - Peguero-Lo Presti voltage (max S + S_V4_) | 0.651 | 0.454 | 0.757 | 0.623 | 0.563 | 0.338 |
| - Cornell voltage (R_avL_ + S_V3_) | 0.622 | 0.427 | 0.684 | 0.522 | 0.567 | 0.353 |
| - Cornell VDP | 0.635 | 0.449 | 0.693 | 0.537 | 0.564 | 0.346 |
| - Sokolow-Lyon voltage (S_V1_ + max R (V5 or V6)) | 0.627 | 0.445 | 0.664 | 0.513 | 0.585 | 0.372 |
| - Gubner-Ungerleider voltage (R_I_ + S_III_) | 0.577 | 0.349 | 0.588 | 0.362 | 0.544 | 0.315 |
| ***ML models*** |  |  |  |  |  |  |
| - Logistic regression^b^ | 0.699 | 0.529 | 0.739 | 0.592 | 0.641 | 0.436 |
| - Random forest^b^ | 0.687 | 0.516 | 0.717 | 0.567 | 0.661 | 0.465 |
| - Light gradient boosted machine^b^ | 0.680 | 0.512 | 0.707 | 0.564 | 0.662 | 0.473 |
| - Residual network^b^ | 0.705 | 0.551 | 0.735 | 0.61 | 0.665 | 0.472 |
| - Multilayered perceptron network^b^ | 0.710 | 0.552 | 0.747 | 0.614 | 0.657 | 0.465 |
| - Convolutional neural network^c^ | 0.668 | 0.497 | 0.697 | 0.527 | 0.661 | 0.483 |

VDP, voltage-duration product; ML, machine learning

* At specificity 0.75

^a^ Logistic regressions

^b^ Input of ECG statistics like QRS duration, heart rate etc. and 30 variational autoencoder latent variables from ECG representative beat and sex

^c^ Input of representative-beat ECG signal (X, Y, Z leads)

**Supplementary table 2D.** Model performance for LVH prediction in population with intraventricular conduction delay (QRS duration >120 ms) validation set.

| **Validation set** | **Combined**  **(n=1,945)** | | **Females**  **(n=814)** | | **Males**  **(n=1,131)** | |
| --- | --- | --- | --- | --- | --- | --- |
|  | **AUROC** | **Sensitivity*** | **AUROC** | **Sensitivity*** | **AUROC** | **Sensitivity*** |
| ***Univariable models^a^*** |  |  |  |  |  |  |
| - QRS duration | 0.677 | 0.51 | 0.742 | 0.544 | 0.611 | 0.46 |
| - Amplitude_QRS-3D_ | 0.499 | 0.275 | 0.553 | 0.281 | 0.550 | 0.336 |
| - Amplitude_QRS-X_ | 0.542 | 0.326 | 0.618 | 0.391 | 0.525 | 0.338 |
| - Amplitude_QRS-Y_ | 0.563 | 0.331 | 0.615 | 0.319 | 0.533 | 0.335 |
| - Amplitude_QRS-Z_ | 0.623 | 0.405 | 0.678 | 0.494 | 0.582 | 0.341 |
| - VTI_QRS-3D_ | 0.613 | 0.371 | 0.597 | 0.293 | 0.608 | 0.409 |
| - VTI_QRS-X_ | 0.575 | 0.343 | 0.602 | 0.311 | 0.572 | 0.378 |
| - VTI_QRS-Y_ | 0.539 | 0.271 | 0.516 | 0.153 | 0.453 | 0.217 |
| - VTI_QRS-Z_ | 0.646 | 0.447 | 0.676 | 0.499 | 0.615 | 0.383 |
| ***Traditional criteria*** |  |  |  |  |  |  |
| - Peguero-Lo Presti voltage (max S + S_V4_) | 0.666 | 0.474 | 0.674 | 0.42 | 0.625 | 0.448 |
| - Cornell voltage (R_avL_ + S_V3_) | 0.664 | 0.476 | 0.73 | 0.521 | 0.583 | 0.384 |
| - Cornell VDP | 0.694 | 0.519 | 0.766 | 0.592 | 0.612 | 0.404 |
| - Sokolow-Lyon voltage (S_V1_ + max R (V5 or V6)) | 0.583 | 0.353 | 0.685 | 0.489 | 0.519 | 0.291 |
| - Gubner-Ungerleider voltage (R_I_ + S_III_) | 0.586 | 0.355 | 0.667 | 0.407 | 0.520 | 0.249 |
| ***ML models*** |  |  |  |  |  |  |
| - Logistic regression^b^ | 0.801 | 0.691 | 0.845 | 0.756 | 0.774 | 0.613 |
| - Random forest^b^ | 0.791 | 0.678 | 0.835 | 0.725 | 0.739 | 0.606 |
| - Light gradient boosted machine^b^ | 0.827 | 0.742 | 0.874 | 0.818 | 0.784 | 0.682 |
| - Residual network^b^ | 0.816 | 0.71 | 0.86 | 0.783 | 0.772 | 0.63 |
| - Multilayered perceptron network^b^ | 0.819 | 0.707 | 0.829 | 0.715 | 0.802 | 0.687 |
| - Convolutional neural network^c^ | 0.801 | 0.696 | 0.821 | 0.733 | 0.782 | 0.661 |

VDP, voltage-duration product; ML, machine learning

* At specificity 0.75

^a^ Logistic regressions

^b^ Input of ECG statistics like QRS duration, heart rate etc. and 30 variational autoencoder latent variables from ECG representative beat and sex

^c^ Input of representative-beat ECG signal (X, Y, Z leads)
